# Developing a social vulnerability index from real-world electronic health records using natural language processing

**DOI:** 10.64898/2026.10.07.26364944

**Authors:** Anna Tirkkonen, Esha Khan, Juho Kaijansinkko, Markus J. Haapanen, Kaisa Koivunen, Anna A. Peltola, Marko Niemelä, Emiel O. Hoogendijk, Jake Lin, Juulia Jylhävä, Mikaela B. von Bonsdorff

**Author notes:** Corresponding author: Anna Tirkkonen.

## Abstract

**Background:** Social vulnerability is associated with adverse health outcomes, but existing social vulnerability indices often rely on cohort-based and self-reported data, potentially limiting their population-level applicability. Therefore, this study aimed to develop an SVI from routinely collected electronic health records (EHRs) using natural language processing (NLP) and examine its association with mortality and healthcare utilization.

**Method:** EHR data from 193,629 individuals aged 35–102 from the Wellbeing Services County of Central Finland between years 2010 and 2023 were used in this study. A 6-item SVI (total score 0-6) was developed from free-text clinical notes using deep-learning-based NLP. SVI scores were calculated at baseline and annually for individuals. Associations between baseline SVI and all-cause mortality and healthcare utilization were assessed over 12-year follow-up using Cox and count models.

**Results:** **T**he mean baseline SVI among all participants was 0.35 (range 0-6). Women had higher SVI (0.39) than men (0.32, p<0.001). SVI increased with age. One unit increase in SVI was associated with a higher risk of mortality (Hazard Ratio (HR) 1.23; 95% Confidence Interval (CI) (1.21-1.24), a higher odds of using healthcare services (Odds Ratio (OR) = 1.20; 95% CI): 1.19–1.21) and a greater healthcare service use (Incidence Rate Ratio (IRR) = 1.07; 95% CI: 1.06–1.07).

**Discussion:** An index capturing multiple domains of social vulnerability can be derived from EHRs using NLP. Higher SVI was associated with increased mortality and healthcare utilization. These findings support the potential of EHR data for assessing social vulnerability and related health risks.

**Highlights:**

- SVI can be derived from free-text EHR data using NLP in an unselected population.
- Higher EHR-derived social vulnerability was associated with mortality and healthcare use.
- EHR data may help identify individuals needing targeted assessment or support

## Introduction

Social vulnerability is a complex phenomenon referring to deficiencies in the quality or quantity of social connections or the level of available support that individuals experience^1^. Disadvantages related to education, income, and social support can accumulate and reinforce each other over the life course, leading to greater social vulnerability and poor health outcomes in later life ^2^. Social vulnerability has been associated with higher risk of frailty, multimorbidity, frailty and premature mortiality,^3,4^ and has also been shown to predict, higher hospital admissions, lower use of preventive healthcare services, including vaccinations and cancer screening, delayed initiation of prenatal care, and poor healthcare experience^5^.

To measure social vulnerability, composite indices have been developed that capture multiple social factors rather than focusing on individual factors such as low educational attainment or limited access to healthcare^6^. These social vulnerability indices (SVIs) integrate socioeconomic, demographic, and environmental domains into a composite measure of vulnerability. By combining multiple dimensions into a single metric, SVIs enable more efficient identification of population at higher risk for adverse health outcomes and inequities compared with analysis based on single factors^3^.

Traditionally, SVIs have been derived from survey or cohort data, which may underrepresent the most vulnerable individuals as those experiencing greater social vulnerability may be less likely to participate in studies^7^. This limit the representativeness and generalisability of findings to the wider population^8^. Artificial intelligence-assisted natural language processing (NLP) offers a scalable approach for extracting information on social determinants of health from unstructured free text in routinely collected clinical records^9,10^. Previous studies have successfully used NLP methods to identify individual social domains^11–13^ but only a limited number have combined these domains into a SVI or examined the association between SVI and adverse health outcomes.

Previous research concerning SVIs has focused primarily on older adults^5^, meaning that the extent to which the SVI captures social vulnerability across the adult life course remains uncertain. Furthermore, SVIs have mainly been developed using data from the US^5^ where healthcare access, mobility-related environmental factors, such as opportunities for walking and reliance on private vehicles, and geographical conditions, including exposure to natural hazards such as hurricanes and earthquakes, differ from those in Nordic countries. These contextual differences may affect the relevance and composition of social vulnerability measures across settings. Together, these gaps highlight the need for new, large-scale approaches to constructing SVIs that can capture social vulnerability more comprehensively across the population.

In this study, we developed a novel SVI based on six individual social vulnerability domains using NLP applied to large-scale longitudinal real-world data that consists of region-wide electronic healthcare records (EHR) from over 200,000 Finnish individuals in the period 2010 to 2023^14^. This index provides a comprehensive understanding of social vulnerability by capturing its dynamic evolution over time and demonstrating its validity in relation to key health outcomes; mortality and healthcare utilization.

The aim of this study was to (1) construct an SVI based on free texts in routinely collected clinical records using NLP; (2) examine how the SVI and its individual domains change over time across different age groups and sexes; and (3) investigate whether the SVI is associated with mortality and healthcare utilization and whether it provides additional predictive value beyond age, sex and multimorbidity.

## 2. Methods

### Study design and sample

The sample encompassed all EHRs from 2010 to 2023, including public in-and outpatient primary, secondary, tertiary, long-term care, and home-care services across the Central Finland Wellbeing Services County^15^. The records included free-text entries of all healthcare professionals, International Classification of Diseases-10 (ICD-10) codes, as well as routine laboratory tests. The free-text entries included, for example, physical examination notes, nursing notes, rehabilitation notes, and patient and family communications^14^. In total, the data included 39.74 million free-text entries from 201 629 individuals aged 35 years and older. For the SVI analysis individuals less than one year of follow-up since the first recorded entry in EHRs were excluded (n=7993), resulting in a final sample size of 193 636 individuals. SVI was calculated for all individuals (Figure 1). Individuals were followed up from entry year until death or the date of the last recorded entry in EHR.

**Figure 1.**
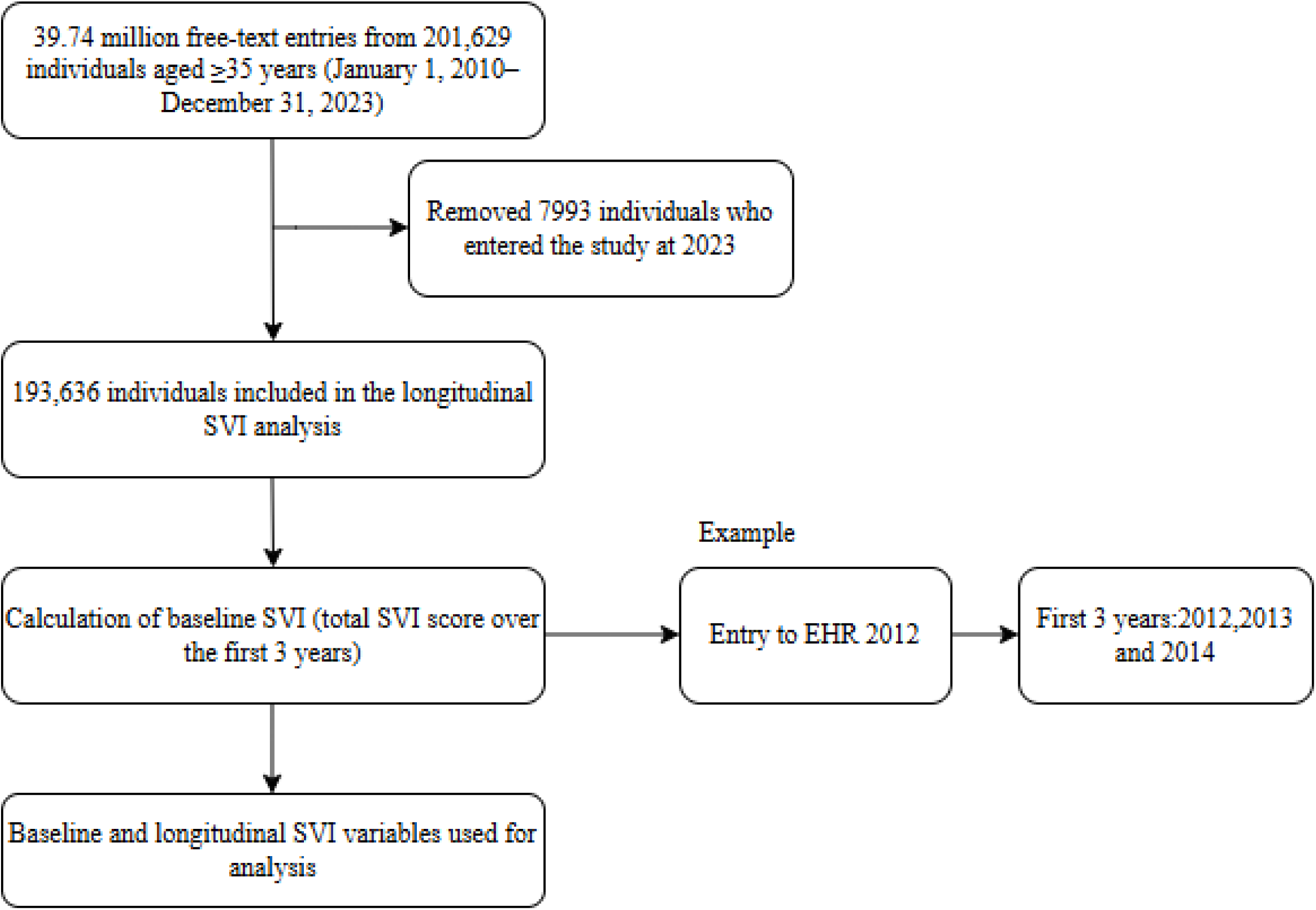
Study flow of the SVI construction

### Social vulnerability index (SVI)

SVI was identified from the EHR data using named entity recognition (NER), based on a fine-tuned FinBERT model (a Finnish-language implementation of BERT architecture), which enabled accurate identification of relevant entities within the text^16^. The NLP NER pipeline has been previously described^14^.

In brief, the SVI comprises six individual social vulnerability domains, including “loneliness”, “not receiving help”, “infrequent contacts with friends or family”, “rarely leaving home”, “living alone”, and “receipt of formal care” (defined as the use of formal care services, such as home care, home help, or home nursing services). The domains were selected based on established dimensions of social vulnerability, including social support, participation, isolation, living arrangements, and reliance on formal care^5,17^ and their relevance to information that can be captured from routine free-texts. Individual social vulnerability domains were considered as binary (yes/no), with each domain contributing one point to the SVI. Higher SVI scores indicated higher levels of social vulnerability, with a total score ranging from 0 to 6.

For “living alone”, “loneliness”, and “receipt of formal care”, a score of 0 indicated a non-vulnerable individual and 1 indicated a vulnerable individual. For “not receiving help and infrequent contacts with friends or family” and “rarely leaving home”, scoring was reversed. For these items, the absence of a positive indication in the EHR text (e.g., receiving help from family or friends or leaving home) was coded as 1 (vulnerable), whereas the presence of a positive indication was coded as 0 (not vulnerable).

### Identification of SVI domains from EHR data

Subsamples for manual labelling of separate domains were extracted from the EHR with relevant keywords/phrases. Relevant texts were labelled by a domain expert and approved by another domain expert. For all domains, positive text entries were labelled to indicate the presence of these domains (e.g., indications of loneliness).

After labelling, the dataset was converted into the BIO tagging format required for the NER model and split into training and validation sets in 90/10 ratio. A separate test set was not used. Using the Hugging Face transformer framework^18^, models were trained and fine-tuned by optimizing the key hyperparameters, learning rate and weight decay. Cross-entropy loss was applied as the default loss function during training. Model performance was evaluated using standard metrics, including precision, recall, and their harmonic mean, the F1 score. The best-performing model for each domain was retained for inference.

Inference was performed using the retained model through the Hugging Face Transformers pipeline API. Candidate texts were first identified using regular expression patterns based on the original domain-specific subsampling keywords. A token-level confidence threshold of 0.95 was applied to retain only high-confidence entity predictions and reduce false positives. SVI calculation.

The potential effect of negated statements (e.g., “patient is not lonely”), a known limitation of NER and a potential source of false-positive predictions, was assessed using a customized approach^19^. Unique labelled text spans were extracted from the original training data and, when possible, manually converted into negated forms (e.g., “lonely” to “not lonely”). The keyword-based search was then repeated across the full dataset using these negated forms, followed by inference on the retrieved texts. Predictions were aggregated at the patient ID level and intersected with the corresponding original SVI domain results. For each domain, the number and proportion of overlapping IDs were quantified to estimate the potential impact of negation-related false-positive predictions on downstream patient-level analyses.

SVI baseline was calculated over the first three years following first EHR. For example, individuals entering the EHR in 2012 had their SVI calculated using data from 2012, 2013, and 2014. Baseline SVI was used to examine the association between SVI and adverse outcomes. Annual SVI values were calculated separately for all individuals and each year of follow-up (2010-2022) using data from that year only. Annual SVI were used to assess changes in SVI over time. Missing values for individual social vulnerability domains were imputed as 0 for years in which the individual had other documented healthcare contacts, but no record of the specific vulnerability indicator. In contrast, years without any patient record entries were retained as missing, as absence of EHRs does not necessarily indicate absence of social vulnerability. For example, the individual may have moved to another area or otherwise no longer been captured in the available EHRs.

### Mortality and Healthcare utilization and covariates

Mortality and healthcare utilization data were obtained from EHRs. Mortality data were linked with national population registers: deaths occurring within healthcare settings were recorded directly in the EHRs, while deaths occurring outside healthcare were transferred to the EHRs from the Digital and Population Data Services Agency. Follow-up for mortality began after the baseline SVI assessment. Healthcare utilization was assessed during the first year following baseline SVI assessment and defined as the total number of primary and secondary care visits.

### Covariates

Information of age in the time of entry (baseline) and sex was drawn from EHRs. Baseline multimorbidity was calculated using the Charlson Comorbidity Index (CCI)^20^, based on ICD-10 codes recorded during SVI baseline period.

### Statistical analyses

Cox proportional hazard models were used to examine the association between baseline SVI and mortality. The base model was fitted with baseline age, sex, and CCI. Then, the SVI baseline was added to the base model, and the model was adjusted for CCI. Model discrimination was assessed with Harrell’s concordance index (C-index) and difference in discrimination between the models was evaluated. Proportional hazards’ assumptions were assessed and confirmed using Schoenfeld residuals. The generalizability of the Cox model was evaluated using a 70:30 train–test split (hold-out validation) with predictive performance assessed on the held-out test set.

The association between baseline SVI and healthcare utilization was analysed using hurdle negative binomial models. A hurdle model estimates the likelihood of any healthcare utilization and the frequency of contacts among users; the binomial component produced odds ratios (ORs) for any use, while the truncated count component produced incidence rate ratios (IRRs) reflecting contact intensity. Model fit for count models was assessed using the Akaike Information Criterion (AIC) and Bayesian Information Criterion (BIC).

Generalizability of the count models was evaluated using a 70:30 train–test split with 10-fold cross-validation, with root mean squared error (RMSE) and mean absolute error (MAE) used to quantify prediction error. Sex-stratified analyses were also performed as a sensitivity analyses. Changes in SVI and individual social vulnerability domains over time were visualized using trend lines of mean SVI values stratified by age and sex.

## Results

### Baseline characteristics

The dataset included 193 636 individuals (51.6% female). The baseline age of all participants was 62.0 year (standard deviation, SD 13.6), ranging from 35 to 100 years. Mean baseline SVI among all individuals was 0.35. Median baseline for all participants as well as for women and men was 0.00. Females had higher SVI at the baseline compared to males (0.39 vs 0.32, p<0.001). Individuals were followed up for median of 11 years (IQR 3 to 14). 43988 individuals died during follow-up period. Baseline characteristics are presented in Table 1.

**Table 1.**
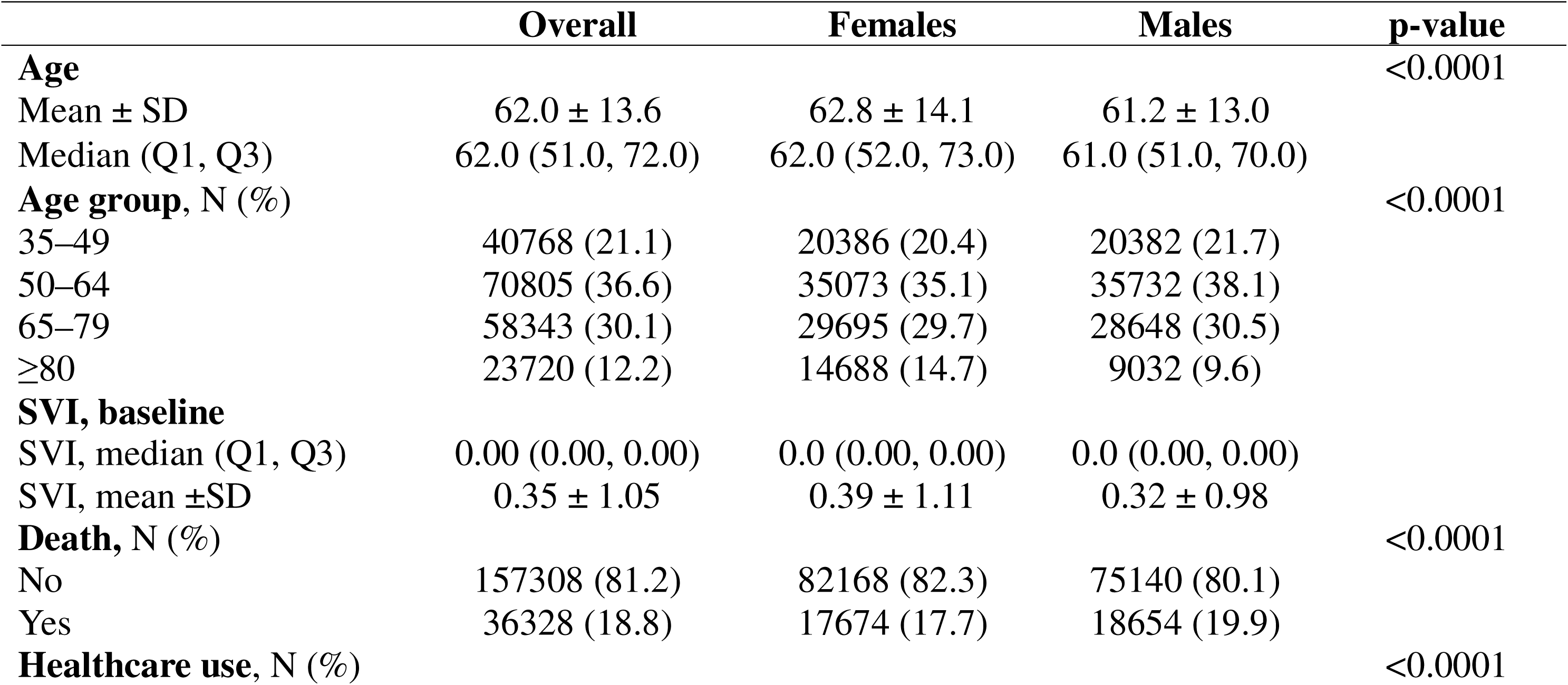
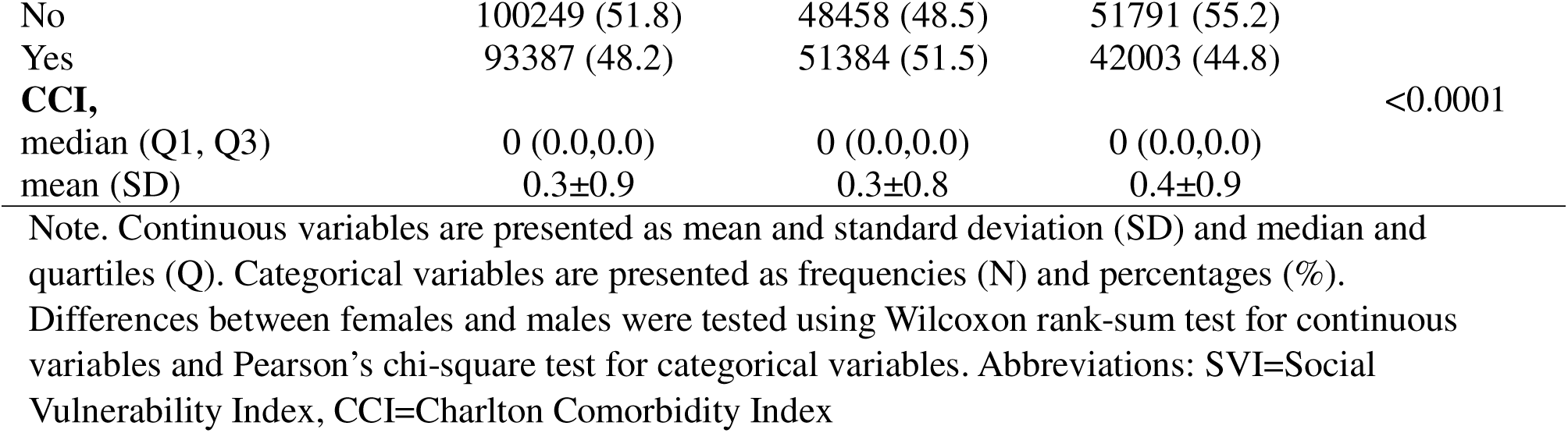
Sample characteristics.

In the NLP NER models F1 score for all items except “not receiving help from family and friends” was above 0.80 indicating excellent model performance. For this item the F1 score was 0.76, indicating good model performance (Supplementary Table 1). The analysis indicated potential negation-related false-positive classifications. Positive-to-negated statement ratios ranged from 8:1 to 12:1 for going outside, rarely leaving home, and loneliness, and from 113:1 to 308:1 for living alone, getting help, and formal care. The proportion of overlapping patient IDs between the original and negated datasets ranged from 33% to 78%, with the highest overlap in domains showing the greatest imbalance between positive and negated statements. Thus, although negation-related false positives were present, their overall impact on patient-level analyses was likely limited.

### Results for SVI

Higher baseline SVI was associated with increased risk for mortality among all individuals (Table 2). Cox models showed that one-unit increase in SVI was associated with an HR of 1.23 (95% confidence interval 1.21-1.24). Complementing base model with SVI increased C-index from 0.84 to 0.85. The relative association was highest in the youngest age group and attenuated with increasing age. In sex-stratified models the HR was slightly higher among men compared to women (1.24 vs 1.20, respectively), but the model demonstrated better discriminative ability among women as reflected by the higher C-index (0.83 vs. 0.88).

**Table 2.**
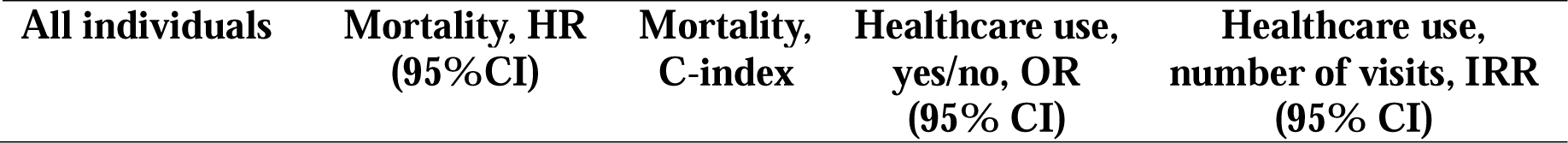
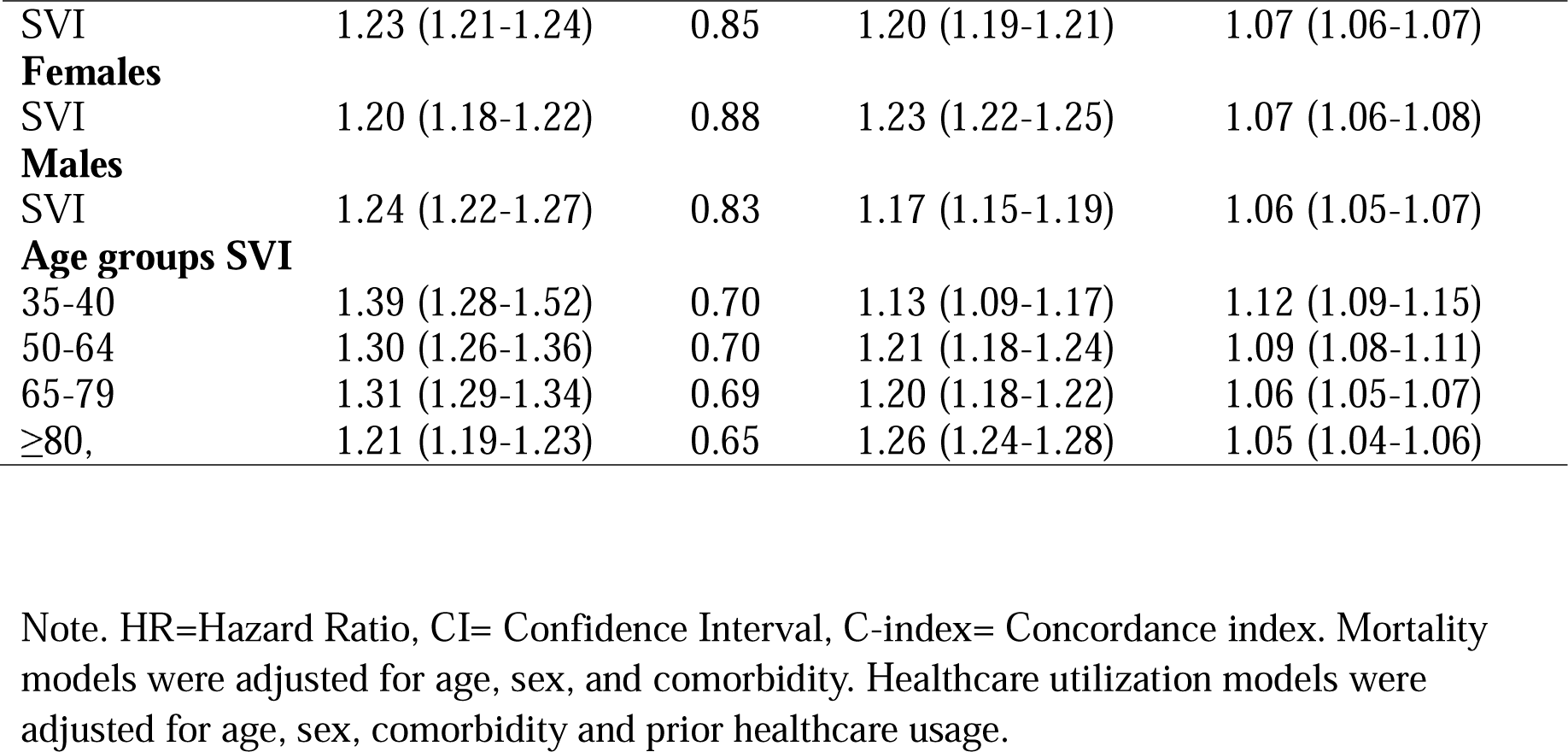
Associations of SVI with mortality and healthcare utilization during follow first year after baseline period using Cox regression and hurdle negative binomial models.

Negative binomial models showed that higher SVI was associated with increased healthcare utilization. A one-unit increase in SVI was associated with 20% higher odds of any healthcare service utilization (OR = 1.20, 95% CI: 1.19–1.21) with the highest relative associations observed in the oldest age group, whereas the estimates were lower in the younger age groups. In sex-stratified models female showed slightly stronger association compared to males (OR 1.23 and 1.17, respectively). Among healthcare services users, a one-unit increase in SVI was associated with a 7% increase in the expected number of healthcare visits (IRR = 1.07, 95% CI: 1.06–1.07). The highest effect was observed among the youngest individuals and weakened with increasing age, whereas no meaningful differences were observed between men and women.

Longitudinally SVI levels varied by age, with higher SVI levels observed among older individuals and the steepest upward pattern among those aged 80 years and older. Younger age groups showed lower SVI levels and more gradual upward patterns during the 12-year follow-up (Figure 2). Although women consistently exhibited higher SVI values than men, SVI showed similar pattern for both sexes during most of the follow-up period.

**Figure 2.**
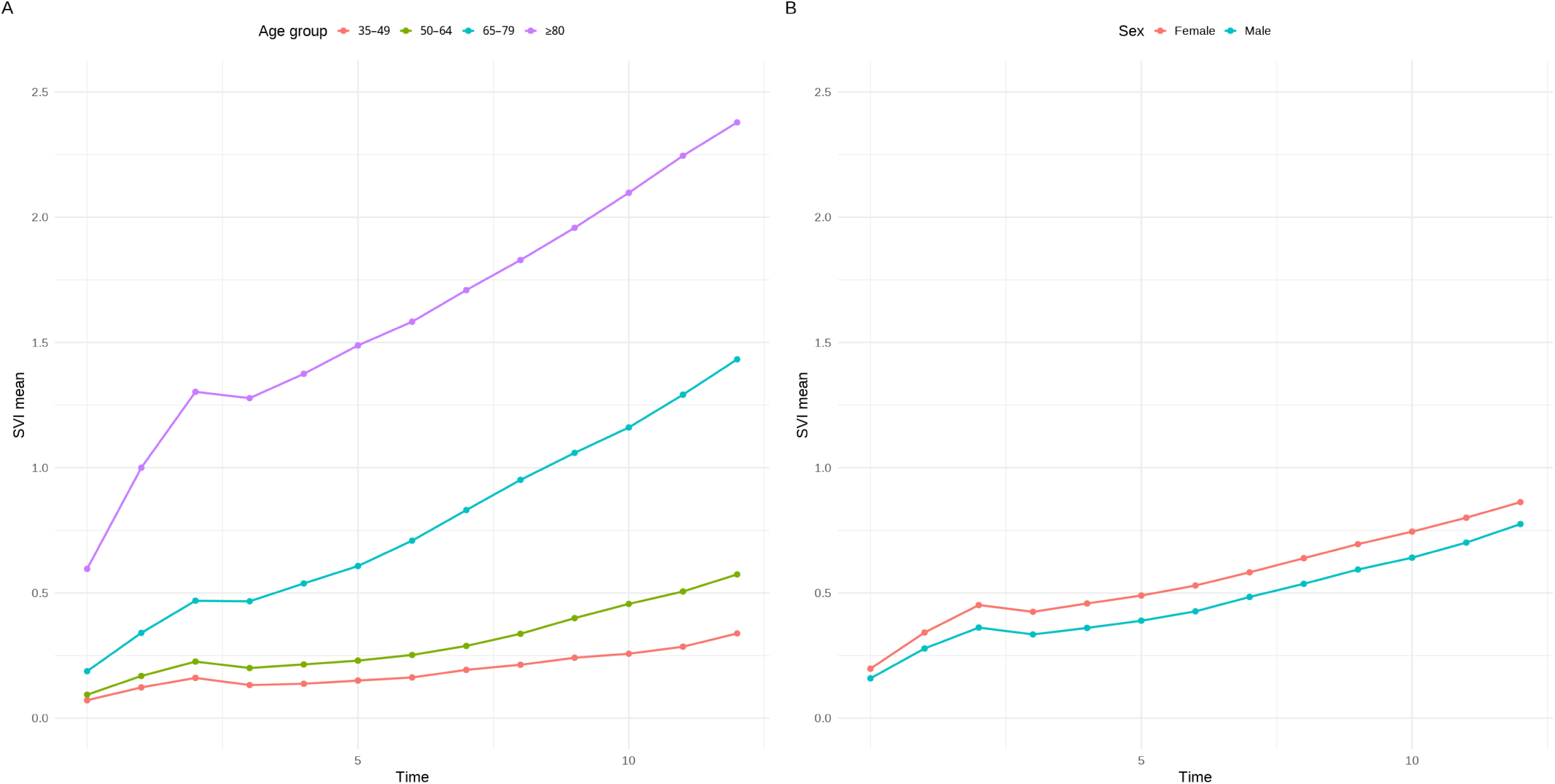
Change in SVI over time by age (panel A) and sex (panel B)

### Results for individual social vulnerability domains

The prevalence of individual social vulnerability domains during follow-up ranged from 4% for “loneliness” to 30% for “infrequent contact with friends or family”. The corresponding prevalences were 7% for “receipt of formal care”, 10% for “living alone”, 28% for “not receiving help from family and friends”, and 29% for “rarely leaving home”. Supplementary Tables 2 and 3 present the distributions and correlations of individual items and the total number of different social vulnerability items observed at least once during follow-up.

Social vulnerability was highest among those aged ≥80 years and progressively lower in younger age groups (Figure 3). Increases over time were most pronounced for “not receiving help from family and friends”, “infrequent contacts with friends or family”, and “rarely leaving home”, whereas “living alone”, “loneliness”, and “receipt of formal care” increased among those aged ≥65 years but showed no significant changes in younger groups. Social vulnerability was slightly higher among females for all items and changes over time were similar between sexes.

**Figure 3.**
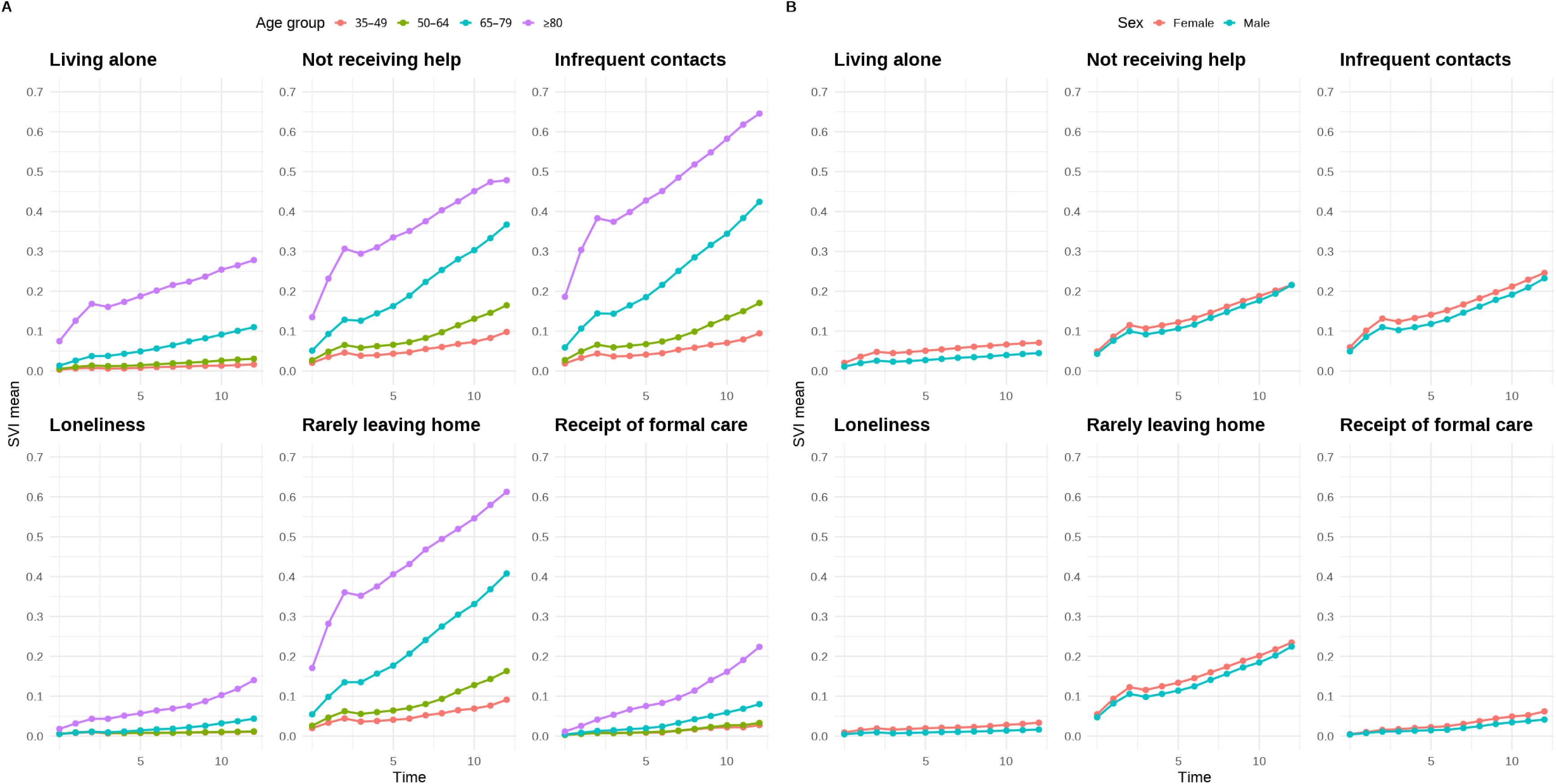
Change in individual social vulnerability items over time by age (panel A) and sex (panel B)

### Generalizability and model fit evaluation

Hold-out validation demonstrated consistent results between the training and test sets (Supplementary Table 4). In training set (n=87618, events= 6344) models C-index was 0.86. In test set (n=37550, events= 2752) C-index was also 0.85 indicating consistent discriminative performance across the training and independent test datasets. Model performance was comparable between the training and test sets. The similarity of RMSE and MAE values between the training and test sets indicates stable model performance and no evidence of overfitting.

## Discussion

In this study, we developed an SVI from free-text EHR entries in a large sample of Finnish individuals aged 35–100 years. We found that SVI increased with age, and higher baseline SVI was associated with both increased risk for mortality and healthcare utilization. Women had slightly higher SVI levels during follow-up, whereas the association between SVI and mortality was stronger among men. These findings suggest that routinely collected EHR data may provide a scalable approach to assessing social vulnerability and its associations with adverse outcomes.

Previous research has shown that social vulnerability increases with age^21,22^, potentially reflecting losses in social networks and participation and age-related declines in physical and cognitive functioning^21,23^. Our findings extend this evidence across a broader adult age range, showing the steepest age gradient among individuals aged ≥80 years and demonstrating that these patterns are observable in real-world clinical data. As previous research has primarily focused on older populations, our broad adult age range extends this evidence by demonstrating an age gradient in social vulnerability from midlife to advanced old age. Moreover, the use of routinely collected EHRs extends previous evidence derived mainly from cohort and survey studies by demonstrating that these age-related patterns are also observable in real-world clinical populations.

We observed sex-specific differences in both SVI levels and its association with mortality. Although higher SVI was associated with increased mortality risk in both sexes, the relative effect of SVI on mortality appeared stronger among men, whereas SVI provided better discrimination of mortality risk among women. Differences in differences in social roles, living arrangements, socioeconomic circumstances, and social integration may contribute to these differences^22^. Several pathways may link social vulnerability to mortality, including adverse health behaviors ^24,25^ chronic stress^26^ and depressive symptoms^27^, and limited social support, which may reduce access to practical assistance and necessary healthcare^28^. However, the mechanisms underlying the association between social vulnerability and mortality, and particularly the observed sex differences, remain unclear.

Our findings indicate that greater social vulnerability was associated with a higher likelihood of healthcare utilization and a higher number of visits among healthcare users, consistent with the systematic review by Valentin et al.^5^ Limited informal support may increase reliance on formal healthcare services, as needs that might otherwise be partly addressed within an individual’s social network may require support from healthcare services. Notably, the association between SVI and higher number of healthcare visits was strongest among the youngest individuals, suggesting that social vulnerability may be particularly relevant for healthcare utilization when it emerges earlier in adulthood. In older age, the higher prevalence of multimorbidity^29^ and greater age-related healthcare needs^30^ may partly explain why the relative contribution of social vulnerability to healthcare utilization was smaller. These findings highlight the potential relevance of social vulnerability for understanding patterns of healthcare use across adulthood and for identifying groups with greater healthcare needs.

Beyond the overall SVI, examining individual social vulnerability domains revealed that the domains showed different patterns over time. Our results showing that individual social vulnerability domains developed differently over time are broadly consistent with previous literature^31–34^. Our results strengthen the evidence that social vulnerability is a multidimensional phenomenon whose individual domains do not evolve uniformly over time. Rather, individual social vulnerability domains may follow distinct trajectories that reflect age-related changes social networks, availability of informal support and decline in physical functioning. Sex differences were generally small, suggesting that sex may play limited role in the development of social vulnerability across domains.

### Strengths and limitations

One of the main strengths of the study was the use of a large real-world dataset, including 200,000 individuals across a broad age range (≥35 years). This captured also the most vulnerable individuals in the population, enhancing the generalizability of the findings. Another main strength was the sophisticated analysis method, which involved analysing free-text data from electronic health records using an NLP-based approach with NER implemented via a fine-tuned Fin BERT model. This approach enables the efficient processing of clinical texts and the context-aware the extraction of social vulnerability-related information from EHRs.

Some limitations affecting generalizability of the findings should be considered. First, the sample was drawn from one healthcare district in Central Finland, although the region includes both urban and rural populations. As employed individuals are usually covered by private occupational health services, they might use public primary healthcare less frequently. Therefore public healthcare records from primary healthcare may underrepresent employed individuals and be slightly socioeconomically skewed. Restricting the sample to individuals with EHR entries may further overrepresent less healthy individuals. Second, the scoring of “not receiving help and infrequent contacts with friends or family” and “rarely leaving home” may slightly overestimate vulnerability because the absence of positive entries was interpreted as vulnerability. However, despite the difference in scoring, all variables showed similar patterns over time. Finally, the NLP models were internally validated using data from the same region as the training data. External validation across other regions, EHR systems, languages, and healthcare settings is therefore warranted.

## Conclusions

We provide evidence that different domains of social vulnerability can be reliably identified from free-text EHRs using NLP, offering a scalable approach for monitoring social risk in clinical populations. In the future, an SVI derived from EHRs may have potential as a tool to support the identification of individuals at increased risk of social vulnerability in health care settings.

## Funding

This work was supported by Research Council of Finland (Grant no: 349335, 349336), Samfundet Folkhälsan i svenska Finland rf, Medicinska Understödsföreningen Liv och Hälsa rf and Yrjö Jahnsson Foundation

## Conflicts of interest

Nothing to declare

## Author contributions

AT, MvB and JJ designed the study, EK, JK and AT performed statistical analysis, AT drafted the initial manuscript, JJ and MvB acquired funding. All authors contributed to interpretation of the findings and commented and edited the final draft.

## Ethical approval

Informed consent and review/or approval by ethics committee were not requires for the study as according to Finnis legislation (Act on the Secondary Use of Health and Social Data 552/2019 by the Ministry of Social Affairs and Health), the individuals included in the study sample were not contacted, and the study did not affect the treatment of the patients. The Human Sciences Ethics Committee of The University of Jyväskylä has certified these conditions pertinent to this study and stated that an ethical review is not required. The study was conducted in accordance with the Declaration of Helsinki.

## Declaration of Generative AI and AI-assisted technologies in the writing process

During the preparation of this work, the authors used generative AI in order to check the grammar and readability of the text. Authors take full responsibility for the content of the publication

## Data Availability

The data used in this study cannot be shared by the authors due to legislative and ethical restrictions; however, all data analyzed in this study are available through an application to the Central Finland Wellbeing Services County

## Notes

### Competing Interest Statement

The authors have declared no competing interest.

### Author Declarations

Informed consent and review/or approval by ethics committee were not requires for the study as according to Finnis legislation (Act on the Secondary Use of Health and Social Data 552/2019 by the Ministry of Social Affairs and Health), the individuals included in the study sample were not contacted, and the study did not affect the treatment of the patients. The Human Sciences Ethics Committee of The University of Jyvaskyla has certified these conditions pertinent to this study and stated that an ethical review is not required. The study was conducted in accordance with the Declaration of Helsinki.

