## Supplementary materials for "Developing a social vulnerability index from real-world electronic health records using natural language processing"

**Supplementary appendix**


| | |
|---|---|
| Page 2 | Supplementary Table 1. Performance metrics of the social vulnerability items identified from free-text items using NLP NER |
| Page 3 | Supplementary Table 2. Number of individuals experiencing social vulnerability at least one time point during follow-up |
| Page 4 | Supplementary Table 3. Correlation between social vulnerability items based on their occurrence during follow-up |
| Page 5 | Supplementary Table 4. Hold-out validation based on model concordance on all-cause mortality |

**Supplementary Table 1. Performance metrics of the social vulnerability items identified from free-text items using NLP NER**

| Category | F1 | Precision | Recall | Evaluation type (strictness) | Learning rate | Weight decay |
|---|---|---|---|---|---|---|
| Not receiving help | 0.759 | 0.66 | 0.89 | ent_type | 1.5E-05 | 0.10000 |
| Rarely leaving home | 0.809 | 0.79 | 0.83 | ent_type | 2.5E-05 | 0.20000 |
| Loneliness | 0.883 | 0.86 | 0.91 | partial | 1.0E-04 | 0.19997 |
| Infrequent contacts with family and friends | 0.928 | 0.88 | 0.98 | ent_type | 4.5E-05 | 0.08000 |
| Receipt of formal care | 0.809 | 0.69 | 0.97 | ent_type | 5.0E-06 | 0.20000 |
| Living alone | 0.894 | 0.82 | 0.99 | ent_type | 5.0E-05 | 0.14000 |

Precision represents the accuracy of positive predictions; Recall indicates the ability to identify all actual positive instances; the F1 score provides the harmonic mean of both precision and recall, serving as a balanced measure of model reliability. Evaluation type defines what counts as a correct prediction relative to the ground truth as established by expert curation and approval; partial evaluation requires that predicted and ground truth spans overlap, regardless of entity type. The ent_type evaluation additionally requires a correct entity type but does not require exact boundary matches. The learning rate controls the size of each weight update during training. Weight decay is a regularization technique that reduces large weights. NLP, natural language processing, NER, named entity recognition.

**Supplementary Table 2**. Number of individuals experiencing social vulnerability at least one time point during follow-up

| | Not receiving help | Rarely leaving home | Loneliness | Infrequent contacts | Receipt of formal care | Living alone |
|---|---|---|---|---|---|---|
| Yes, N (%) | 53 368 (28) | 55358 (29) | 7711(4) | 56856 (29) | 12769 (7) | 18439 (10) |
| No, N (%) | 140268 (72) | 138278 (71) | 185925 (96) | 136780 (71) | 180867 (93) | 175197 (90) |

**Supplementary Table 3**. **Correlation between social vulnerability items based on their occurrence during follow-up**

| | | Not receiving help | Rarely leaving home | Loneliness | Infrequent contacts | Receipt of formal care | Living alone |
|---|---|---|---|---|---|---|---|
| **Not receiving help** | *r* | 1 | .907 | .310 | .924 | .410 | .484 |
| | *p* | | <0.001 | <0.001 | <0.001 | <0.001 | <0.001 |
| **Rarely leaving home** | *r* | .907 | 1 | .313 | .949 | .414 | .501 |
| | *p* | <0.001 | | <0.001 | <0.001 | <0.001 | <0.001 |
| **Loneliness** | *r* | .310 | .313 | 1 | .306 | .237 | .310 |
| | *p* | <0.001 | <0.001 | | <0.001 | <0.001 | <0.001 |
| **Infrequent contacts** | *r* | .924 | .949 | .306 | 1 | .410 | .496 |
| | *p* | <0.001 | <0.001 | <0.001 | | <0.001 | <0.001 |
| **Receipt of formal care** | *r* | .410 | .414 | .237 | .410 | 1 | .276 |
| | *p* | <0.001 | <0.001 | <0.001 | <0.001 | | |
| **Living alone** | *r* | .484 | .501 | .310 | .496 | .276 | 1 |
| | *p* | <0.001 | <0.001 | <0.001 | <0.001 | <0.001 | <0.001 |

Note: Each item was coded as 1 if the vulnerability was identified at least once during follow-up and 0 otherwise

**Supplementary Table 4. Hold-out validation based on model concordance on all-cause mortality**

| Mortality | N | Events | C-index | SE |
|---|---|---|---|---|
| Training set | 87 618 | 6344 | 0.86 | 0.004 |
| Test set | 37 550 | 2752 | 0.85 | 0.006 |
